# Risk factors for mortality in patients with Coronavirus disease 2019 (COVID-19) infection: a systematic review and meta-analysis of observational studies

**DOI:** 10.1101/2020.04.09.20056291

**Authors:** Mohammad Parohan, Sajad Yaghoubi, Asal Seraji, Mohammad Hassan Javanbakht, Payam Sarraf, Mahmoud Djalali

## Abstract

**Purpose:** Coronavirus disease 2019 (COVID-19) is an emerging disease that was first reported in Wuhan city, the capital of Hubei province in China, and has subsequently spread worldwide. Risk factors for mortality have not been well summarized. Current meta-analysis of retrospective cohort studies was done to summarize available findings on the association between age, gender, comorbidities and risk of death from COVID-19 infection.

**Methods:** Online databases including Web of Science, PubMed, Scopus, Cochrane Library and Google scholar were searched to detect relevant publications up to 1 May 2020, using relevant keywords. To pool data, random-effects model was used. Furthermore, sensitivity analysis and publication bias test were also done.

**Results:** In total, 14 studies with 29,909 COVID-19 infected patients and 1,445 cases of death were included in the current meta-analysis. Significant associations were found between older age (≥65 vs <65 years old) (pooled ORs=4.59, 95% CIs=2.61-8.04, p<0.001), gender (male vs female) (pooled ORs=1.50, 95% CIs=1.06-2.12, p=0.021) and risk of death from COVID-19 infection. In addition, hypertension (pooled ORs=2.70, 95% CIs= 1.40-5.24, p=0.003), cardiovascular diseases (CVDs) (pooled ORs=3.72, 95% CIs=1.77-7.83, p=0.001), diabetes (pooled ORs=2.41, 95% CIs=1.05-5.51, p=0.037), chronic obstructive pulmonary disease (COPD) (pooled ORs=3.53, 95% CIs=1.79-6.96, p<0.001) and cancer (pooled ORs=3.04, 95% CIs=1.80-5.14, p<0.001), were associated with higher risk of mortality.

**Conclusion:** Older age (≥65 years old), male gender, hypertension, CVDs, diabetes, COPD and malignancies were associated with greater risk of death from COVID-19 infection. These findings could help clinicians to identify patients with poor prognosis at an early stage.

## Introduction

In December, 2019, severe acute respiratory syndrome coronavirus 2 (SARS-CoV-2; previously known as 2019-nCoV) was first reported in Wuhan city, the capital of Hubei province in China, and has subsequently spread to other regions of China and 210 countries and territories [1-3]. SARS-CoV-2, which belongs to a unique clade of the sarbecovirus subgenus of the Orthocoronavirinae subfamily [4], was later designated coronavirus disease 2019 (COVID-19) in February, 2020, by World Health Organization.

Patients with COVID-19 present primarily with fever, dry cough and fatigue or myalgia [5]. Although most patients with COVID-19 are thought to have a favorable prognosis, older patients and those with chronic diseases may have worse outcomes [6]. Patients with chronic underlying conditions may develop viral pneumonia, dyspnea and hypoxemia within 1week after onset of the disease, which may progress to respiratory or end-organ failure and even death [7].

Several studies have reported the clinical characteristics and risk factors associated with death in patients with COVID-19 pneumonia [2,6,8-11]. We aimed to systematically review the present evidences on the association between age, gender, hypertension, diabetes, chronic obstructive pulmonary disease (COPD), cardiovascular diseases (CVDs) and risk of death from COVID-19 infection, and to summarize the available findings in a meta-analysis.

## Methods

### Study protocol

The present systematic review and meta-analysis were planned, conducted and reported in adherence to the Preferred Reporting Items for Systematic Reviews and Meta-Analyses (PRISMA) guidelines [12].

### Search strategy

We performed a literature search using the online databases of Web of Science, PubMed, Scopus, Cochrane Library and Google scholar for relevant publications up to 1 May 2020. The following medical subject headings (MeSH) and non-MeSH keywords were used in our search strategy: (“novel coronavirus” OR “severe acute respiratory syndrome coronavirus 2” OR “SARS-CoV-2” OR “COVID-19” OR “2019-nCoV”) AND (“death” OR “mortality” OR “survival” OR “fatal outcome”). Literature search was done by two independent researchers (MP and SY). We also searched the reference lists of the relevant articles to identify missed studies. No restriction was applied on language and time of publication. To facilitate the screening process of articles from databases, all literature searches were downloaded into an EndNote library (version X8, Thomson Reuters, Philadelphia, USA). The search strategy is presented in detail in Supplementary Table 1.

### Eligibility Criteria

In our meta-analysis, eligible articles were included if they met the following inclusion criteria: (1) all studies assessing the association between age, gender, comorbidities and mortality risk from COVID-19 infection as the major outcomes of interest; (2) observational studies with retrospective design; (3) those that reported hazard ratios (HRs), odds ratios (ORs) or relative risks (RRs) along with 95% confidence intervals (CIs) for the relationship between risk factors and COVID-19 mortality. Review articles, expert opinion articles, theses and books were excluded.

### Data extraction and assessment for study quality

Two investigators (MP and AS) extracted the following data from the included studies: study design, the first author’s name, the publication year, age and gender of patients, sample size, exposure (risk factors), outcome (the risk of mortality), exposure and outcome assessment methods, most adjusted risk estimate (HRs, ORs, RRs) with 95% confidence intervals and adjusted confounding variables.

The Newcastle–Ottawa Scale (NOS) was used for assessing the quality of included retrospective cohort studies based on the following three major components: selection of the study patients, adjustment for potential confounding variables and assessment of outcome [13]. Based on this scale, a maximum of nine points can be awarded to each study. In the present study, articles with the NOS score of ≥ 5 were considered as high quality publications.

### Statistical analysis

We used HRs, ORs, and RRs (and their 95% confidence intervals) reported for the association between risk factors and mortality from COVID-19 infection, to calculate log RRs and their standard errors (SEs). Then, the overall effect size for mortality in relation to risk factors was calculated using random-effects model. For examining the between-study heterogeneity, we performed the Cochran’s Q test (I^2^ ≥ 50% were considered between-study heterogeneity) [14]. To identify potential sources of heterogeneity, we did subgroup analysis according to the predefined criteria as follows: age (≥65 vs. <65), gender (male vs. female), hypertension (yes vs. no), diabetes (yes vs. no), COPD (yes vs. no) and CVDs (yes vs. no). In addition to the main analysis, we carried out sensitivity analysis to find if the overall estimate depended on the effect size from a single study. Assessing the publication bias was done by the formal test of Egger [15]. All statistical analyses were conducted using Stata, version 14.0 (Stata Corp, College Station, TX, USA). P-values were considered significant at level of < 0.05.

## Results

### Search results

In our initial search, we found 143 papers. Of these, 15 duplicates, 17 non-English, 26 non-human, 46 reviews and 17 studies that did not fulfill our eligibility criteria were excluded, leaving 22 papers for further evaluation. Out of remaining 22 papers, 8 were excluded because of the following reason: did not report HRs, ORs or RRs with 95% CIs. Finally, we included 14 retrospective studies in the current systematic review and meta-analysis (Figure 1).

**Figure 1.**
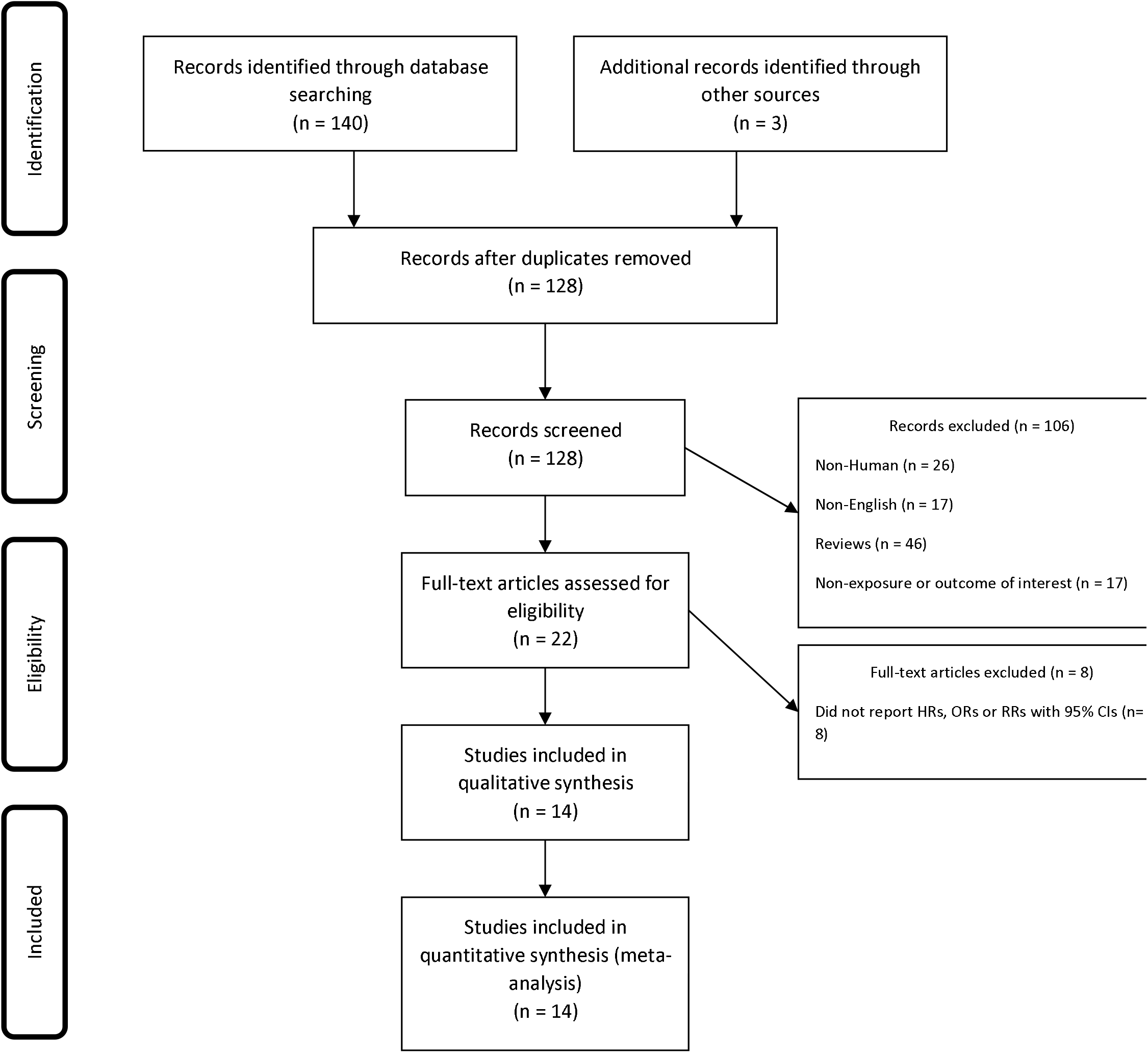
Flow chart of study selection.

### Study characteristics

Twelve studies were conducted in China [2,6,8-11,16-21], one in Italy [22] and one in Iran [23]. The sample size of studies varied from 172 to 20812 patients (mean age, 59.6 years). All studies used real-time reverse transcriptase–polymerase chain reaction (RT-PCR) to confirm COVID-19 infection [2,6,8-11,16-23]. The NOS scores ranged between 5 to 8.

### Demographic characteristics and risk of death from COVID-19

In the meta-analysis of 6 effect sizes, obtained from 6 studies [6,9,10,16,17,22] (3,088 patients and 344 cases of death), we found that older age (≥65 vs <65 years old) was associated with a 459% (over fourfold) increased risk of COVID-19 mortality (pooled ORs=4.59, 95% CIs=2.61-8.04, p<0.001, I^2^=67.1%, p_heterogeneity_=0.010) (Figure 2). Combining 5 effect sizes from 5 studies [6,8-10,23] revealed significant association between male gender and COVID-19 mortality (pooled ORs=1.50, 95% CIs=1.06-2.12, p=0.021, I^2^=76.3%, p_heterogeneity_=0.002) (Figure 2).

**Figure 2.**
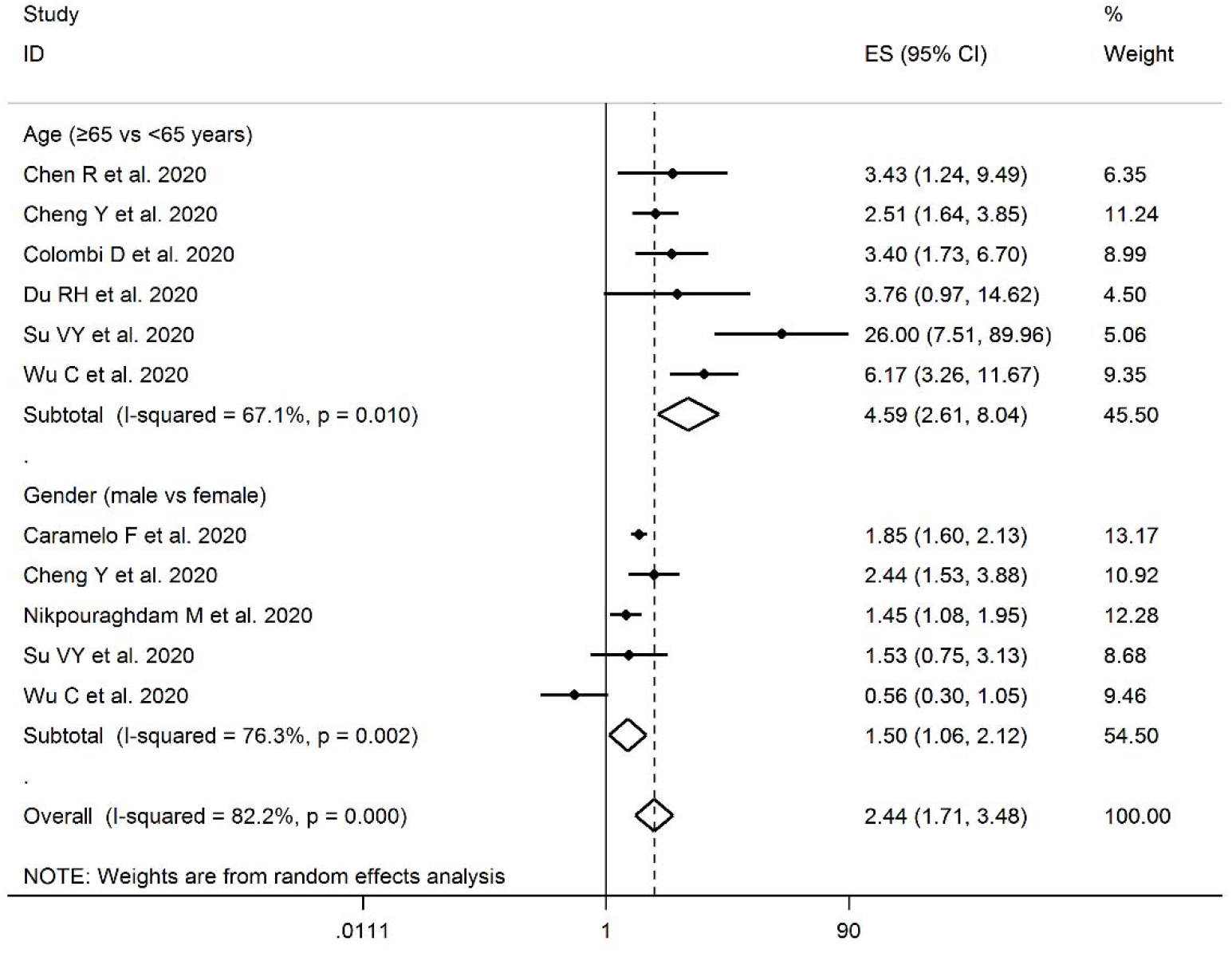
Forest plot for the association between age, gender and risk of mortality from COVID-19 using random-effects model.

### Comorbidities and risk of death from COVID-19

Totally, 38 effect sizes from 12 studies [2,6,8,10,11,16-22] with a total of 26,235 patients and 1,117 cases of death were extracted for the association between comorbidities and COVID-19 mortality. Combining the reported estimates, we found a significant positive association between hypertension (pooled ORs=2.70, 95% CIs= 1.40-5.24, p=0.003, I^2^=92.6%, p_heterogeneity_<0.001) (Figure 3), cardiovascular diseases (CVDs) (pooled ORs=3.72, 95% CIs=1.77-7.83, p=0.001, I^2^=89.1%, p_heterogeneity_<0.001) (Figure 3), diabetes (pooled ORs=2.41, 95% CIs=1.05-5.51, p=0.037, I^2^=93.6%, p_heterogeneity_<0.001) (Figure 4), chronic obstructive pulmonary disease (COPD) (pooled ORs=3.53, 95% CIs=1.79-6.96, p<0.001, I^2^=72.2%, p_heterogeneity_=0.001) (Figure 4), cancer (pooled ORs=3.04, 95% CIs=1.80-5.14, p<0.001, I^2^=41.6%, p_heterogeneity_=0.114) (Figure 4), and risk of death from COVID-19. We found that hypertension, CVDs, diabetes, COPD and cancer were associated with 270% (over twofold), 372% (over threefold), 241% (over twofold), 353% (over threefold) and 304% (over threefold) higher risk of COVID-19 mortality, respectively.

**Figure 3.**
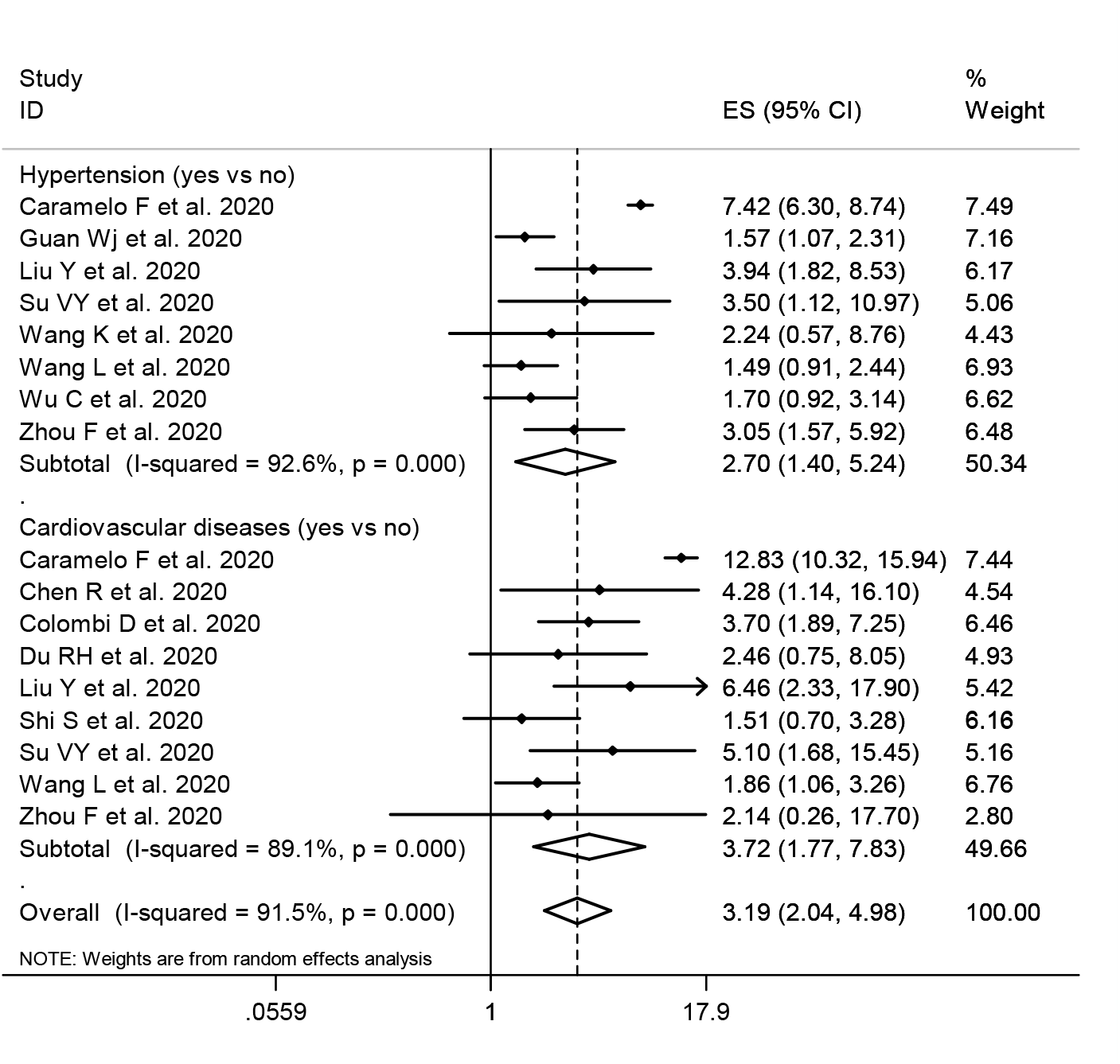
Forest plot for the association between hypertension, cardiovascular diseases and risk of mortality from COVID-19 using random-effects model.

**Figure 4.**
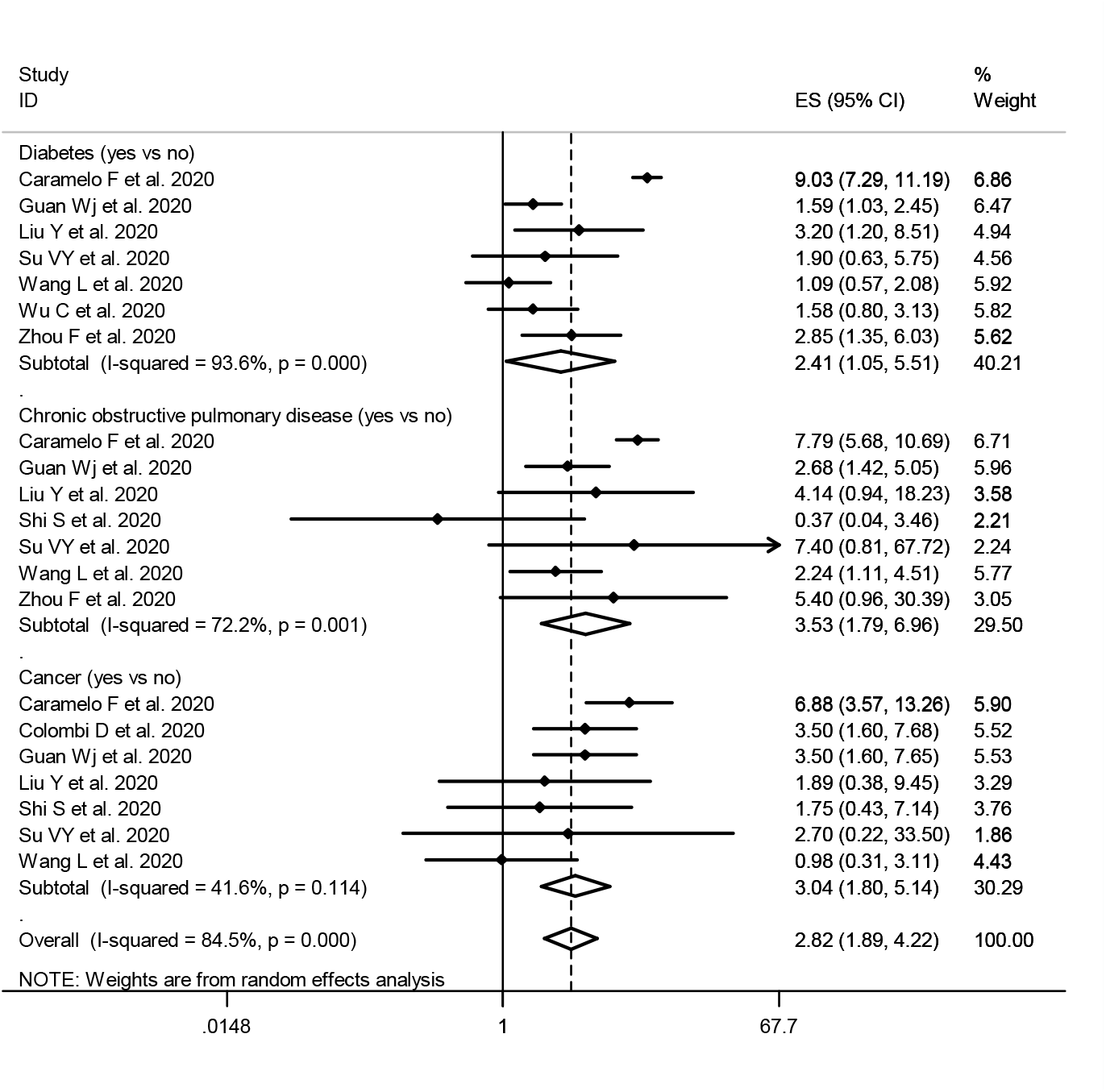
Forest plot for the association between diabetes, chronic obstructive pulmonary disease, cancer and risk of mortality from COVID-19 using random-effects model.

### Sensitivity analysis and publication bias

Findings from sensitivity analysis showed that overall estimates on the association of demographic characteristics and comorbidities with COVID-19 mortality did not depend on a single study (Supplementary Figures 1, 2). Furthermore, based on the results of Egger’s test (age: p=0.185, gender: p=0.388, hypertension: P=0.065, CVDs: P=0.068, diabetes: P=0.117, COPD: P=0.178, and cancer: p=0.054), we found no evidence of publication bias.

## Discussion

Findings from the current systematic review and meta-analysis supported the hypothesis that older age (≥65 years old), male gender, hypertension, CVDs, diabetes, COPD and cancer were associated with higher risk of mortality from COVID-19 infection.

Our findings are partially in agreement with previous narrative review [24]. Previously, older age has been reported as an important risk factor for mortality in SARS and Middle East respiratory syndrome (MERS) [25,26]. The current meta-analysis confirmed that increased age (≥65 years old) was associated with death in COVID-19 patients. The age-dependent defects in B-cell and T-cell function and the excess production of type 2 cytokines could lead to prolonged proinflammatory responses and deficiency in control of viral replication, potentially leading to poor outcome [27]. In addition, elderly patients may have other risk factors, such as sarcopenia and comorbidities [11].

Previous studies suggested that COVID-19 infection is more likely to affect older males with comorbidities, and can result in fatal respiratory diseases such as acute respiratory disease syndrome [10,28]. Interestingly, SARS and MERS also infected more males compared to females [29,30]. Differences in the levels and type of circulating sex hormones in males and females might influence the susceptibility of COVID-19 infection. Previous study showed that sex hormones modulate the responses of adaptive and innate immunity [31].

The other risk factors related to death include hypertension, CVDs, diabetes, respiratory system disease and malignancy. A previous study showed that hypertension and diabetes are more prevalent in patients with severe MERS infection [30]. Similarly, the mortality rate of influenza was significantly higher in patients with hypertension, metabolic disease, CVDs and respiratory system disease [32]. Previous studies reported that high protein expression of angiotensin converting enzyme 2 (ACE2) receptor, the receptor for COVID-19, in specific organs correlated with organ failures in SARS patients [33-36]. It has been shown that circulating ACE2 levels are higher in male patients with hypertension, diabetes and CVDs [37,38]. Therefore, male patients with these comorbidities may be more prone to die from COVID-19 infection because of the high expression of ACE2 receptor, though further research on the mechanism is needed.

The pathogenesis of COVID-19 is still not completely understood. Cytokine storm is thought to play an important role in disease severity [39]. Neutrophilia was found in both the lung and peripheral blood of patients with SARS [40,41]. The severity of lung damage correlated with higher numbers of neutrophils and macrophages in the peripheral blood and extensive pulmonary infiltration of these cells in patients with MERS [42-44]. Neutrophils are the main source of cytokines and chemokines. The generation of cytokine storm can lead to acute respiratory distress syndrome, which is a leading cause of death in patients with SARS and MERS [44,45]. This may explain the positive association between high fever and acute respiratory distress syndrome found at the early stages of COVID-19 infection [6].

The present study has some limitations. First, interpretation of our meta-analysis findings might be limited by the small sample size. However, by including studies conducted in different designated hospitals for COVID-19, we believe our findings are representative of cases in Wuhan, China. Second, our meta-analysis did not include data such as smoking history and body mass index, which are potential risk factors for disease severity and mortality.

## Conclusion

Older age (≥65 years old), male gender, hypertension, CVDs, diabetes, COPD and cancer were associated with greater risk of death from COVID-19 infection. The results of the present meta-analysis could help clinicians to identify high risk groups that should receive off-label medications or invasive supportive care, as soon as possible.

## Data Availability

All data are publicly available.

## Author contributions

MP, SY and MD had the idea for the article, MP, AS, MHJ, PS performed the literature search and data analysis, and MP, SY and MD drafted and critically revised the work.

## Compliance with ethical standards

## Ethical approval

Not applicable.

## Informed consent

Not applicable.

## Availability of data and materials

All data generated or analyzed during this study are included in this manuscript.

---

**Table 1.**
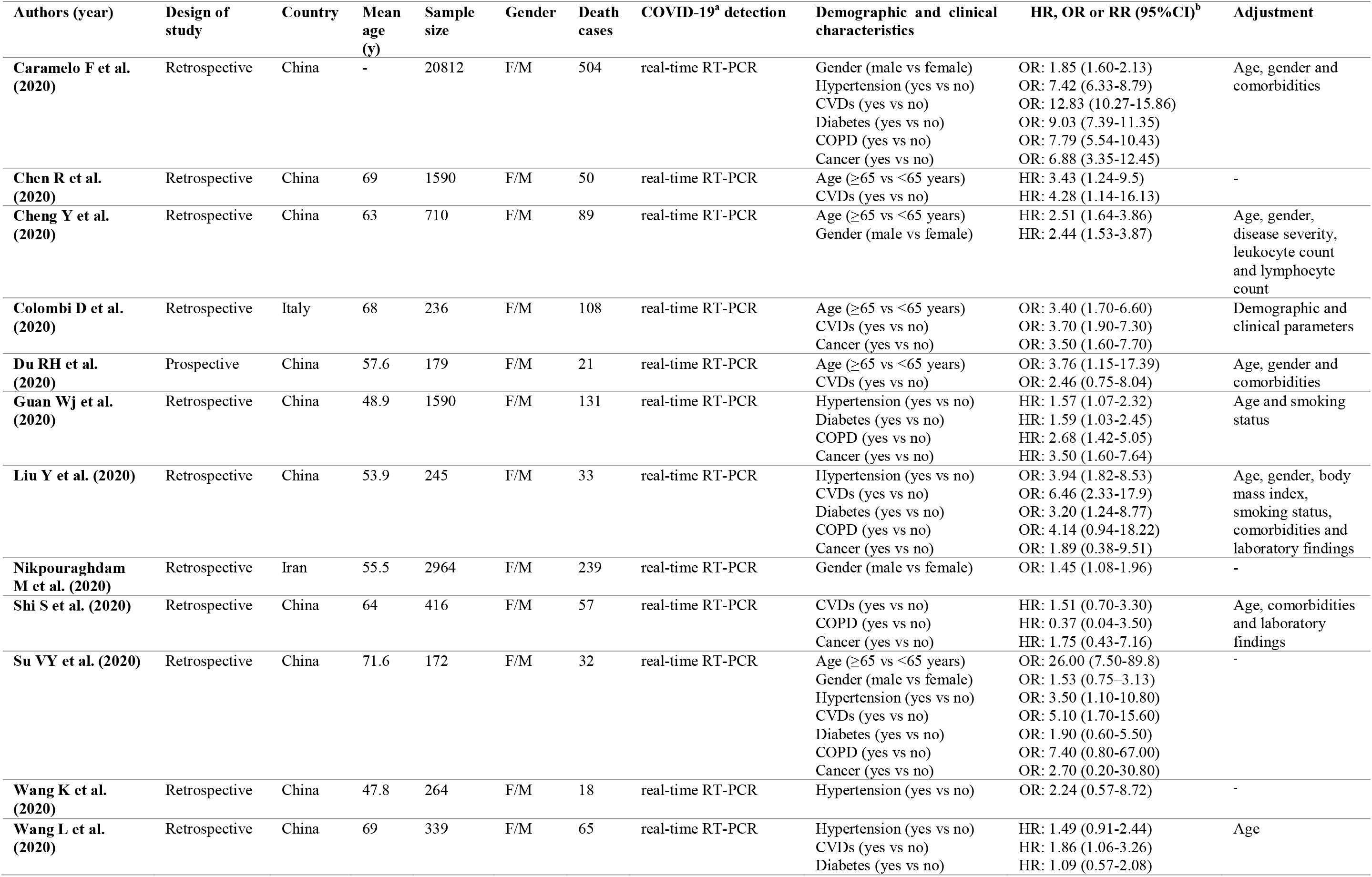
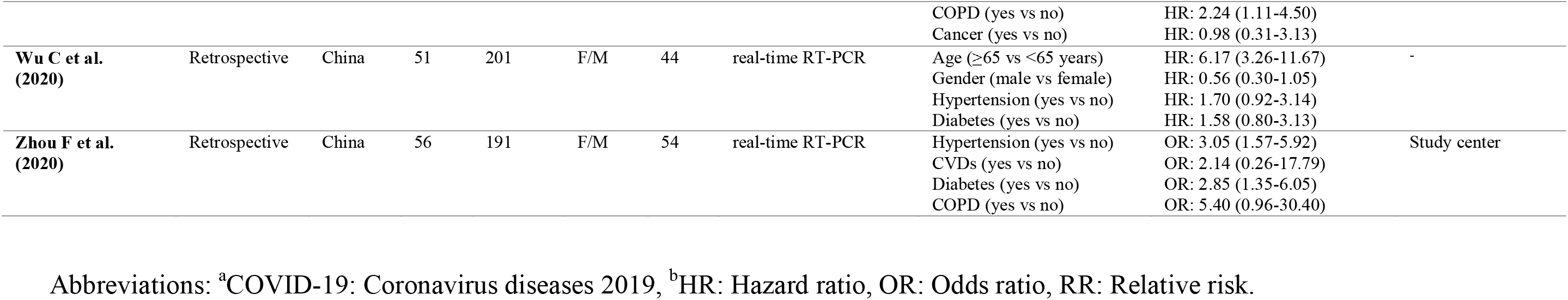
Characteristics of studies included in the meta-analysis.

**Supplementary table 1.** Systematic literature review search terms and strategy.

|  |
| --- |
| **Search terms for PubMed** |
| #1 (“COVID-19”[Mesh] OR “COVID-19”[Title/Abstract] OR “severe acute respiratory syndrome coronavirus 2”[Mesh] OR “severe acute respiratory syndrome coronavirus 2”[Title/Abstract] OR “SARS-CoV-2”[Title/Abstract] OR “novel coronavirus” [Title/Abstract] OR “2019-nCoV”[Title/Abstract]) |
| #2 (“Death”[Mesh] OR “Death”[Title/Abstract] OR “Mortality”[Mesh] OR “Mortality”[Title/Abstract] OR “Survival”[Mesh] OR “Survival”[Title/Abstract] OR “Fatal Outcome”[Mesh] OR “Fatal Outcome”[Title/Abstract]) |
| #1 AND #2 |
| **Search terms for Scopus** |
| #1 (“COVID-19” OR “severe acute respiratory syndrome coronavirus 2” OR “SARS-CoV-2” OR “novel coronavirus” OR “2019-nCoV”): Title/Abstract/Keyword |
| #2 (“death” OR “mortality” OR “survival” OR “fatal outcome”): Title/Abstract/Keyword |
| #1 AND #2 |
| **Search terms for Web of Science** |
| #1. TS = (“COVID-19” OR “severe acute respiratory syndrome coronavirus 2” OR “SARS-CoV-2” OR “novel coronavirus” OR “2019-nCoV”) |
| #2. TS = (“death” OR “mortality” OR “survival” OR “fatal outcome”) |
| #3. #1 AND #2 |
| **Search terms for Cochrane Library** |
| #1 (“COVID-19” OR “severe acute respiratory syndrome coronavirus 2” OR “SARS-CoV-2” OR “novel coronavirus” OR “2019-nCoV”): Title/Abstract/Keyword |
| #2 (“death” OR “mortality” OR “survival” OR “fatal outcome”): Title/Abstract/Keyword |
| #1 AND #2 |

**Supplementary Figure 1.**
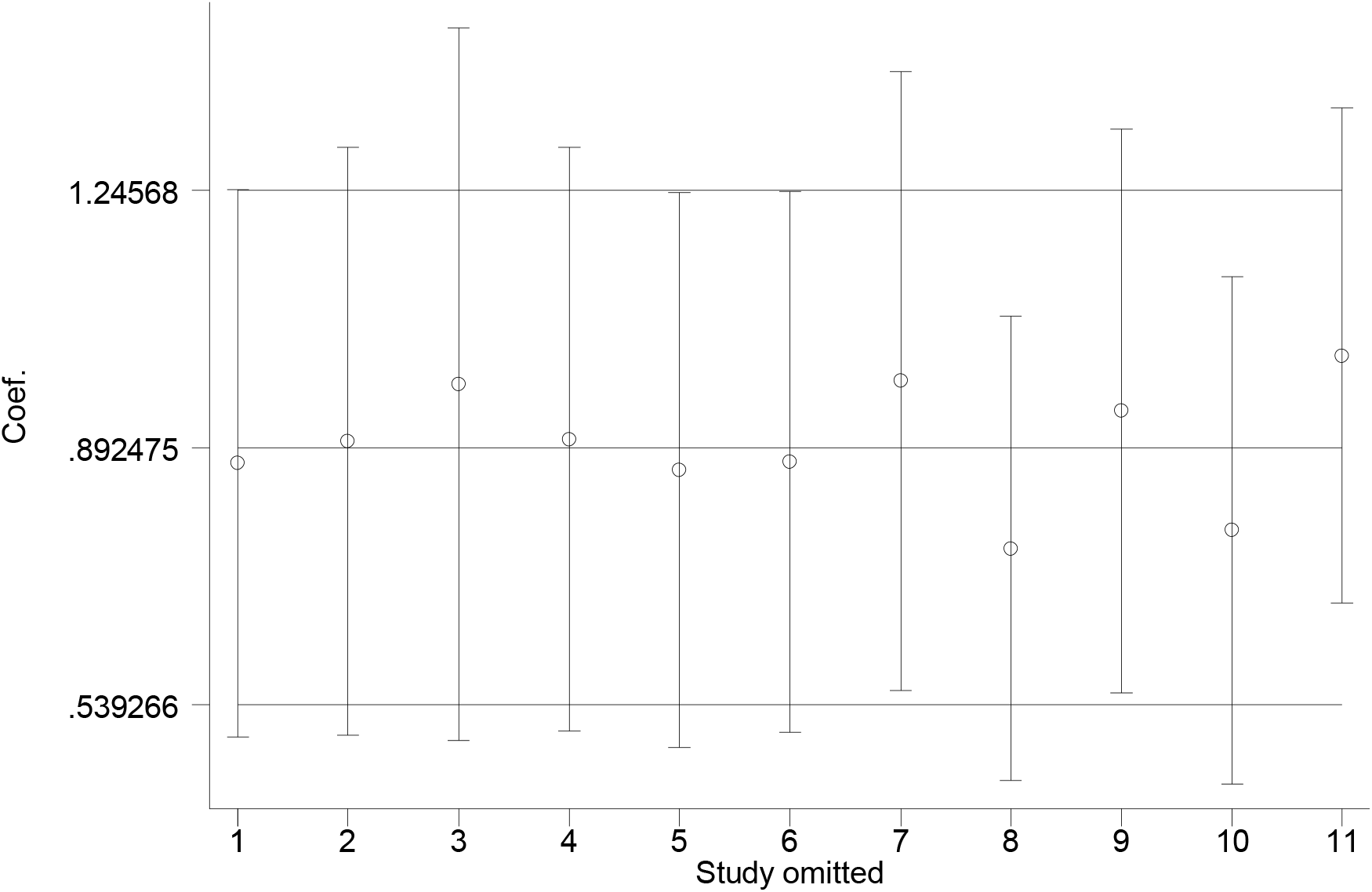
Sensitivity analysis graph for the association between demographic characteristics and risk of death from COVID-19 infection. The results of the sensitivity analysis showed that no study had an obvious influence on the outcomes of this meta-analysis.

**Supplementary Figure 2.**
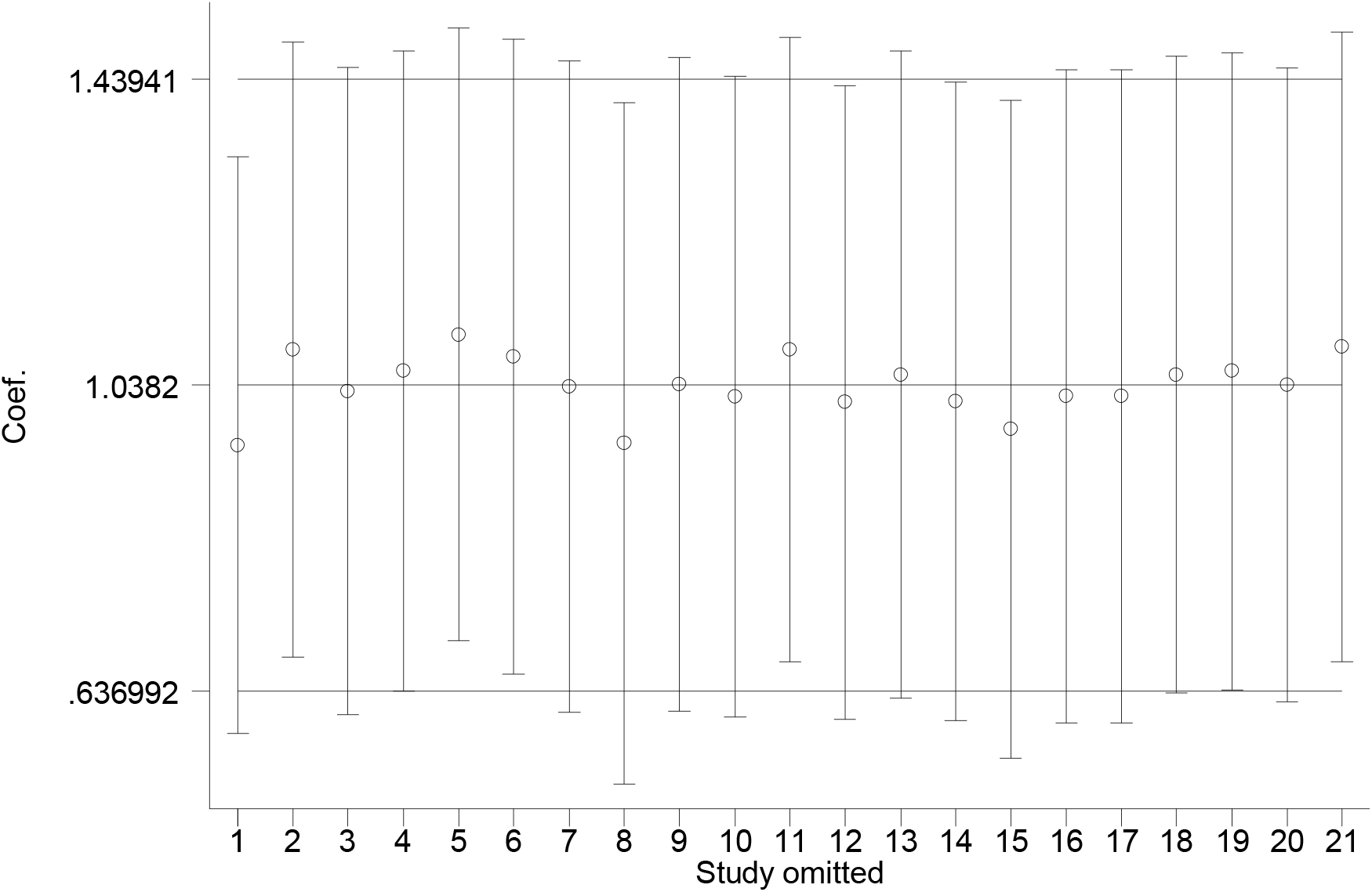
Sensitivity analysis graph for the association between comorbidities and risk of death from COVID-19 infection. The results of the sensitivity analysis showed that no study had an obvious influence on the outcomes of this meta-analysis.

PRISMA 2009 Checklist
| Section/topic | # | Checklist item | Reported on page # |
| --- | --- | --- | --- |
| **TITLE** |  |  |  |
| Title | 1 | Identify the report as a systematic review, meta-analysis, or both. | 1 |
| **ABSTRACT** |  |  |  |
| Structured summary | 2 | Provide a structured summary including, as applicable: background; objectives; data sources; study eligibility criteria, participants, and interventions; study appraisal and synthesis methods; results; limitations; conclusions and implications of key findings; systematic review registration number. | 2 |
| **INTRODUCTION** |  |  |  |
| Rationale | 3 | Describe the rationale for the review in the context of what is already known. | 4 |
| Objectives | 4 | Provide an explicit statement of questions being addressed with reference to participants, interventions, comparisons, outcomes, and study design (PICOS). | 4 |
| **METHODS** |  |  |  |
| Protocol and registration | 5 | Indicate if a review protocol exists, if and where it can be accessed (e.g., Web address), and, if available, provide registration information including registration number. | 5 |
| Eligibility criteria | 6 | Specify study characteristics (e.g., PICOS, length of follow-up) and report characteristics (e.g., years considered, language, publication status) used as criteria for eligibility, giving rationale. | 5 |
| Information sources | 7 | Describe all information sources (e.g., databases with dates of coverage, contact with study authors to identify additional studies) in the search and date last searched. | 5 |
| Search | 8 | Present full electronic search strategy for at least one database, including any limits used, such that it could be repeated. | 5 |
| Study selection | 9 | State the process for selecting studies (i.e., screening, eligibility, included in systematic review, and, if applicable, included in the meta-analysis). | 5 |
| Data collection process | 10 | Describe method of data extraction from reports (e.g., piloted forms, independently, in duplicate) and any processes for obtaining and confirming data from investigators. | 6 |
| Data items | 11 | List and define all variables for which data were sought (e.g., PICOS, funding sources) and any assumptions and simplifications made. | 6 |
| Risk of bias in individual studies | 12 | Describe methods used for assessing risk of bias of individual studies (including specification of whether this was done at the study or outcome level), and how this information is to be used in any data synthesis. | 6 |
| Summary measures | 13 | State the principal summary measures (e.g., risk ratio, difference in means). | 6,7 |
| Synthesis of results | 14 | Describe the methods of handling data and combining results of studies, if done, including measures of consistency (e.g., $I^2$) for each meta-analysis. | 6,7 |

PRISMA 2009 Checklist
| Section/topic | # | Checklist item | Reported on page # |
| --- | --- | --- | --- |
| Risk of bias across studies | 15 | Specify any assessment of risk of bias that may affect the cumulative evidence (e.g., publication bias, selective reporting within studies). | 7 |
| Additional analyses | 16 | Describe methods of additional analyses (e.g., sensitivity or subgroup analyses, meta-regression), if done, indicating which were pre-specified. | 7 |
| **RESULTS** |  |  |  |
| Study selection | 17 | Give numbers of studies screened, assessed for eligibility, and included in the review, with reasons for exclusions at each stage, ideally with a flow diagram. | 7 |
| Study characteristics | 18 | For each study, present characteristics for which data were extracted (e.g., study size, PICOS, follow-up period) and provide the citations. | 7 |
| Risk of bias within studies | 19 | Present data on risk of bias of each study and, if available, any outcome level assessment (see item 12). | 7 |
| Results of individual studies | 20 | For all outcomes considered (benefits or harms), present, for each study: (a) simple summary data for each intervention group (b) effect estimates and confidence intervals, ideally with a forest plot. | 8,9 |
| Synthesis of results | 21 | Present results of each meta-analysis done, including confidence intervals and measures of consistency. | 8,9 |
| Risk of bias across studies | 22 | Present results of any assessment of risk of bias across studies (see Item 15). | 9 |
| Additional analysis | 23 | Give results of additional analyses, if done (e.g., sensitivity or subgroup analyses, meta-regression [see Item 16]). | 9 |
| **DISCUSSION** |  |  |  |
| Summary of evidence | 24 | Summarize the main findings including the strength of evidence for each main outcome; consider their relevance to key groups (e.g., healthcare providers, users, and policy makers). | 9-11 |
| Limitations | 25 | Discuss limitations at study and outcome level (e.g., risk of bias), and at review-level (e.g., incomplete retrieval of identified research, reporting bias). | 11 |
| Conclusions | 26 | Provide a general interpretation of the results in the context of other evidence, and implications for future research. | 11 |
| **FUNDING** |  |  |  |
| Funding | 27 | Describe sources of funding for the systematic review and other support (e.g., supply of data); role of funders for the systematic review. | 11 |

From: Moher D, Liberati A, Tetzlaff J, Altman DG, The PRISMA Group (2009). Preferred Reporting Items for Systematic Reviews and Meta-Analyses: The PRISMA Statement. PLoS Med 6(7): e1000097. doi:10.1371/journal.pmed1000097
For more information, visit: www.prisma-statement.org.

